# Offline Reinforcement Learning for Out-of-Distribution ICU Sepsis Decision Support

**DOI:** 10.64898/2026.08.22.26361090

**Authors:** Emad Arasteh, Maryam S. Mirian, Maryam Tavakol

## Abstract

Offline reinforcement learning (RL) provides a promising framework for learning and evaluating treatment policies from logged clinical data, particularly in sequential decision-making settings where prospective exploration would be unsafe. In ICU sepsis management, however, it remains unclear whether offline RL policies retain stable behavior under increasingly severe out-of-distribution (OOD) patient cohorts. In this paper, we evaluate standard offline RL methods on three severity-enriched OOD test mixtures from the MIMIC-III benchmark dataset to determine whether offline policies retain a stable, action-sensitive decision-support signal. Under the shared learned-dynamics off-policy evaluation (OPE) protocol, as the severe-OOD ratio increases from 25% to 75%, observed clinical survival declines from 67% to 49%, while the best offline method in each mixture receives model-predicted terminal survival values of 87%, 86%, and 85%, respectively. Because observed clinical survival and model-predicted terminal survival are different quantities, this contrast suggests a stable model-based decision-support signal under severity shift. We further present a secondary physiological stabilization analysis using an episode-level physiological stabilization score (EPSS), a heuristic summary of whether selected physiological variables move in favorable directions during follow-up. In this analysis, model-generated rollouts under offline policies receive higher EPSS values than matched logged clinical trajectories for several physiological components. Together, these results support learned-dynamics OPE as a useful severity-OOD stress test for offline RL policies in ICU sepsis, while leaving prospective and causal validation as necessary next steps.

## 1 Introduction

Sepsis management in intensive care is a critical sequential decision-making problem, where clinicians repeatedly adapt interventions as patient physiology evolves under uncertainty. This structure naturally motivates reinforcement learning (RL) approaches for learning treatment policies from logged ICU data, as demonstrated by [5]. However, as prospective exploration is ethically constrained in such settings, the learned policies must be evaluated carefully, particularly with respect to behavior-policy support and off-policy bias [3]. The concerns become even more acute under distribution shift: a policy that appears reasonable near the training distribution may behave differently for the patients with severe health conditions, who are often underrepresented in the training data but are clinically the most consequential. Offline RL is designed for fixed logged datasets and includes methods that constrain or regularize actions toward behavior-policy support, thereby mitigating extrapolation error under distribution shift [8]. This distinction is important in healthcare: support-aware offline RL can make model-based evaluation more stable, but it does not remove the need for cautious off-policy evaluation (OPE), calibration, and clinical validation.

In this paper, we study standard offline reinforcement learning methods for ICU sepsis decision support under severity-induced distribution shift. We construct out-of-distribution (OOD) test mixtures with increasing proportions of severe held-out trajectories from the MIMIC-III benchmark dataset [4] and evaluate whether offline RL policies retain a stronger model-predicted terminal survival return than the historical clinical survival rate under this stress test. In addition, we provide a physiological stabilization analysis based on model-generated rollouts, showing whether offline-policy trajectories exhibit complementary stabilization signals beyond the terminal survival endpoint. Our results should be interpreted as model-based decision-support signals under severity shift, rather than as causal evidence of clinical benefit or superiority over observed treatment decisions.

Our contributions are threefold. First, we evaluate standard offline RL policies under severity-induced OOD shift by progressively increasing the fraction of severe held-out ICU sepsis trajectories in the test cohort. Second, we report observed logged clinical survival and model-predicted terminal survival return as separate quantities, so that their numerical contrast is interpreted only as a model-dependent OPE diagnostic and not as evidence of causal treatment benefit. Third, we add a secondary physiological stabilization analysis using an episode-level physiological stabilization score (EPSS) computed on model-generated rollouts, providing a complementary diagnostic of whether selected physiological variables move in favorable directions under the learned evaluator.

## 2 The Evaluation Framework

This section describes our evaluation protocol to study offline RL under severity-induced distribution shift in ICU sepsis management. We first define the OOD test cohorts, then introduce the policy-learning methods, the learned-dynamics evaluator, and the experimental endpoints.

### 2.1 Data and OOD Splits

We use ICU-sepsis trajectories derived from the MIMIC-III dataset [4]. Each ICU time step is represented as a one-step transition containing the current continuous-valued patient state vector, a discrete treatment-action bin, a reward signal, the next patient state, and labels indicating whether the episode ended and what terminal outcome was observed. We construct three severity-induced evaluation sets using the patient severity scores available in the data. Policies are learned on the less-severe training cohort and evaluated on held-out test sets with increasing severe-case enrichment. We define trajectory severity using the mean `shock_severity_proxy` value across ICU time bins. For each random seed, severity cutoffs are estimated from the training data, and severe-OOD test episodes are drawn from held-out severe trajectories. Thus, the 25%, 50%, and 75% settings indicate the fraction of severe held-out episodes in the test mixture, not severity-score intervals. Increasing the severe-OOD ratio therefore makes the evaluation cohort progressively harder by replacing more near-distribution held-out samples with severe held-out cases.

The pipeline separates policy learning from model-based evaluation. In the first part, the logged training split is used to train the policy-learning methods in Section 2.2, where each method learns a treatment policy that maps the current patient state to an action. In the second part, the training set is used to learn a dynamics ensemble that is used only for off-policy evaluation, where its architecture, training targets, and rollout use are described in Section 2.3. The offline RL policies do not use this dynamics ensemble during training. At test time, observed clinical survival and EPSS for the logged clinical trajectories are computed directly from the held-out historical trajectories, whereas policy-dependent survival and EPSS are computed by rolling each trained policy through the shared learned-dynamics evaluator.

Furthermore, we compute the observed clinical survival rate from the logged terminal outcomes across *N* episodes in each selected cohort,

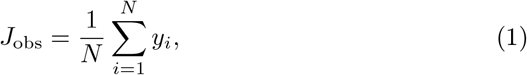

where *y*_*i*_ *∈ {*0, 1*}* denotes the recorded final survival outcome for episode *i*. This quantity is fixed by the logged data and does not depend on any learned policy.

### 2.2 Offline RL Methods

We compare a set of standard offline RL methods under the same fixed-dataset training framework to assess how different strategies behave under the same severity-OOD stress test. All methods are trained only on logged trajectories for policy learning and do not interact with the environment during training or evaluation. The learned policies hence give a treatment strategy that maps the current ICU state to a discrete action.

– **Fitted Q-Iteration (FQI) [1]**. A standard value-based technique, using fitted Bellman updates to learn a Q-function from the fixed dataset with a neural-network function approximator.
– **Conservative Q-Learning (CQL) [7]**. Learns conservative Q-values by penalizing unsupported actions, reducing overestimation in OOD regions.
– **FQI with a CQL-style penalty (FQI+CQL)**. We include a custom hybrid approach that augments the FQI Bellman loss with a CQL-style conservative regularizer using *α* = 0.25.
– **Batch-Constrained Q-Learning (BCQ) [2]**. Restricts policy updates by the logged behavior policy, reducing extrapolation error on OOD actions.
– **Implicit Q-Learning (IQL) [6]**. Avoids any direct maximization over unseen actions and extracts a policy via advantage-weighted regression using an expectile value baseline.
– **Critic-Regularized Regression (CRR) [10]**. Learns a policy by weighted regression toward dataset actions, while using critic-based advantages to prioritize higher-value actions.
– **Advantage-Weighted Actor-Critic (AWAC) [9]**. Adapts advantage-weighted policy updates to favor high-value actions from the data. Although AWAC is originally proposed for offline pretraining followed by online fine-tuning, we use it as a fixed-dataset baseline without any online interaction.

The methods above therefore provide the policies being compared. In the next section, we present how the dynamics ensemble is learned and used for model-based off-policy evaluation. For a given split and seed, the same dynamics ensemble is shared across all evaluated policies, providing a fixed evaluator. Because the dynamics model conditions on both the current state and the selected action, different policies can generate different trajectories from the same initial patient state, which makes the model-based endpoint action-sensitive.

### 2.3 Dynamics Ensemble for Model-Based Off-Policy Evaluation

After policy training, we use a separately learned dynamics ensemble for model-based off-policy evaluation. The ensemble is trained on logged training transitions of the form (*s*_*t*_, *a*_*t*_, *r*_*t*_, *s*_*t*+1_, *d*_*t*_) and is not used to update or optimize the offline policies in Section 2.2. The same evaluator (after training step is finished) is used for all policies within a given split and seed, so differences in model-generated outcomes arise from the actions selected by the evaluated policies rather than from using different dynamics models for different methods.

Architecturally, each ensemble member is a feed-forward multilayer perceptron to model the one-step dynamics. The input is the normalized current ICU state concatenated with a one-hot encoding of the discrete treatment action. This input is passed through two fully connected hidden layers with 256 units and ReLU activations, followed by separate output heads for the normalized next-state change, one-step reward, termination probability, and raw survival probability. During training, logged patient trajectories from the training split are flattened into one-step transitions (*s*_*t*_, *a*_*t*_, *r*_*t*_, *s*_*t*+1_, *d*_*t*_). The model is trained to predict the next-state delta *s*_*t*+1_ *− s*_*t*_, reward, done label, and survival target using supervised losses. Multiple models with the same architecture are trained as an ensemble using bootstrap resampling of the training transitions, and their averaged predictions define the rollout dynamics used for evaluation.

During evaluation, rollouts initiate from held-out states in the selected severity-OOD evaluation subset. For a fixed split, seed, and OOD ratio, the same selected held-out episodes are reused across the compared policies. At each step, the evaluated policy maps the current model state to a treatment action. The learned dynamics ensemble then takes this state-action pair as input and predicts the next state together with a per-step termination probability. The policy is then queried again on the predicted next state, and this alternating process continues until the cumulative termination probability reaches the chosen *p*_done_ threshold or the maximum rollout horizon is reached. Thus, the policy determines the sequence of actions, while the dynamics model determines the model-generated state transitions. The resulting trajectories are action-sensitive model predictions, not observed counterfactual patient trajectories.

### 2.4 Rollout-Stopping Threshold *p*_**done**_

The threshold *p*_done_ is used only in the OPE procedure, but not as a training hyperparameter or as an output of the evaluated policy. At each rollout step, the learned dynamics ensemble predicts a per-step termination probability 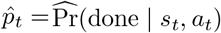 for the current model-generated state and policy-selected action. This value describes the model-predicted probability that the rollout terminates at step *t*. We do not stop the rollout based only on this one-step probability. Instead, we accumulate the probability of termination over the rollout, and define the survival mass *S*_*t*_ and the cumulative termination probability

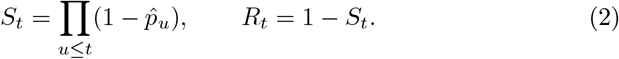

Here, *S*_*t*_ represents the remaining probability mass of not yet being terminated by step *t*, while *R*_*t*_ represents the accumulated probability that termination has occurred by step *t*. The rollout stops when *R*_*t*_ reaches the chosen thresh-old *p*_done_, or when the maximum horizon *H*_max_ = 100 is reached. Larger values of *p*_done_ require more accumulated termination probability before stopping, and therefore usually produce longer model-generated rollouts. Because there is no single clinically observed value of *p*_done_ for these model-generated rollouts, we evaluate several thresholds, *p*_done_ ∈ {0.3, 0.5, 0.7, 0.9}, and report the full sweep to assess whether the qualitative results are sensitive to this stopping threshold.

### 2.5 Model-Predicted Terminal Survival Rate

This subsection defines the policy-dependent survival endpoint used in the model-based policy evaluation. For a selected severity-OOD evaluation cohort with *N* held-out episodes, we compute the model-predicted terminal survival return for policy *π* under rollout-stopping threshold *p*_done_ as

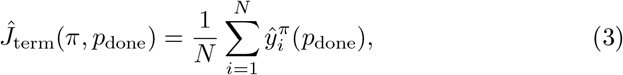

where *i* indexes held-out episodes in the selected evaluation cohort, and the summation is therefore over *N* episodes, not over rollout time steps. In the saved evaluation implementation, 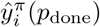 is the calibrated terminal survival probability assigned by a separate survival evaluator to the final model-generated state reached by policy *π* under the learned-dynamics rollout.

Although the learned dynamics ensemble also outputs a raw survival-head probability, we use the calibrated survival evaluator applied to the final model-generated state, rather than the raw survival head directly. Consequently, *Ĵ*_term_ is action-sensitive because it depends on the actions chosen by the evaluated policy, and model-dependent because the trajectory is generated by the learned-dynamics evaluator. By contrast, *J*_obs_ summarizes observed outcomes under historical clinical practice and the true environment dynamics. The contrast between *Ĵ*_term_ and *J*_obs_ should therefore be interpreted as a model-based decision-support signal, not as a causal treatment effect or evidence that the learned policy would improve real patient survival if deployed clinically [3, 8].

### 2.6 Physiological Stabilization Score

We compute a heuristic episode-level physiological stabilization score (EPSS) on the same held-out severity-OOD evaluation sets. For each combination of episode, policy, mixture, seed, and *p*_done_, we compare the initial state with the state at step *h ∈ {*0, 1, 3, 5, 10, 20*}*, where each horizon corresponds to one 4-hour ICU bin. Each physiological component is scored using a direction-corrected rule: (+1) if the variable improves beyond a prespecified tolerance, (0) if it remains stable within tolerance, and (-1) if it worsens. EPSS is then computed as the unweighted mean over the component scores. Table 1 outlines the corresponding state variables, improvement directions, target ranges where applicable, and tolerances used in the score computation. Offline-policy EPSS is computed from model-generated rollouts under the actions selected by the policy. Logged-clinical EPSS is computed from the observed patient state sequence on the matched held-out episode. Table 2 reports horizon-specific availability for the *p*_done_ = 0.9 in the EPSS experiment. Unavailable horizons are excluded from the analysis.

**Table 1.**
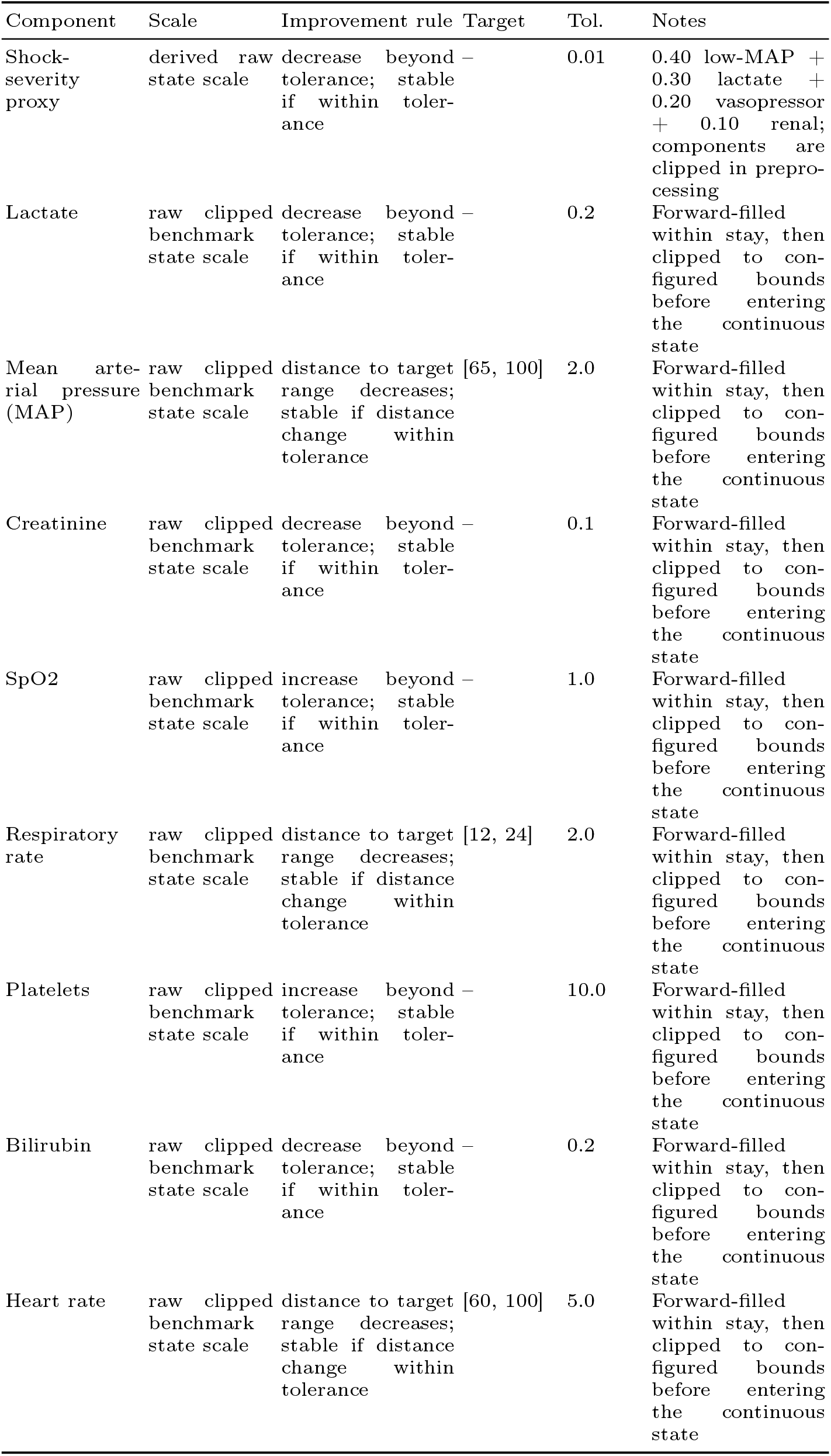
EPSS components definitions used in the physiological stabilization analysis.

**Table 2.** Horizon availability for the CQL-versus-logged EPSS comparison at *p*_done_ = 0.9. Counts are aggregated over the 40 seed-specific test mixtures. For each severity-OOD level, the matched baseline contains 5,120 episode-seed observations.

| Severe-OD | $h$ | Hours | Baseline $N$ | Logged $N$ (%) | CQL rollout $N$ (%) |
| --- | --- | --- | --- | --- | --- |
| 25% | 0 | 0 | 5120 | 5120 (100.0) | 5120 (100.0) |
| 25% | 1 | 4 | 5120 | 5120 (100.0) | 5120 (100.0) |
| 25% | 3 | 12 | 5120 | 5120 (100.0) | 5120 (100.0) |
| 25% | 5 | 20 | 5120 | 5016 (98.0) | 5120 (100.0) |
| 25% | 10 | 40 | 5120 | 4396 (85.9) | 5120 (100.0) |
| 25% | 20 | 80 | 5120 | 3064 (59.8) | 4960 (96.9) |
| 50% | 0 | 0 | 5120 | 5120 (100.0) | 5120 (100.0) |
| 50% | 1 | 4 | 5120 | 5120 (100.0) | 5120 (100.0) |
| 50% | 3 | 12 | 5120 | 5120 (100.0) | 5120 (100.0) |
| 50% | 5 | 20 | 5120 | 4988 (97.4) | 5120 (100.0) |
| 50% | 10 | 40 | 5120 | 4323 (84.4) | 5120 (100.0) |
| 50% | 20 | 80 | 5120 | 3085 (60.3) | 4984 (97.3) |
| 75% | 0 | 0 | 5120 | 5120 (100.0) | 5120 (100.0) |
| 75% | 1 | 4 | 5120 | 5120 (100.0) | 5120 (100.0) |
| 75% | 3 | 12 | 5120 | 5120 (100.0) | 5120 (100.0) |
| 75% | 5 | 20 | 5120 | 4935 (96.4) | 5120 (100.0) |
| 75% | 10 | 40 | 5120 | 4233 (82.7) | 5120 (100.0) |
| 75% | 20 | 80 | 5120 | 2880 (56.2) | 5007 (97.8) |

## 3 Empirical Results

### 3.1 Severity-OOD Survival Evaluation

We begin by examining the observed clinical survival rate in the three severity-OOD test mixtures. As the proportion of severe held-out trajectories increases, the observed clinical survival rate falls from *J*_obs_ = 0.673 at 25% severe-OOD to 0.588 at 50% and 0.488 at 75%. This monotone decline confirms that the constructed test mixtures represent increasingly severe held-out patient cohorts and provides the clinical reference curve shown in Figure 1(b).

**Fig. 1.**
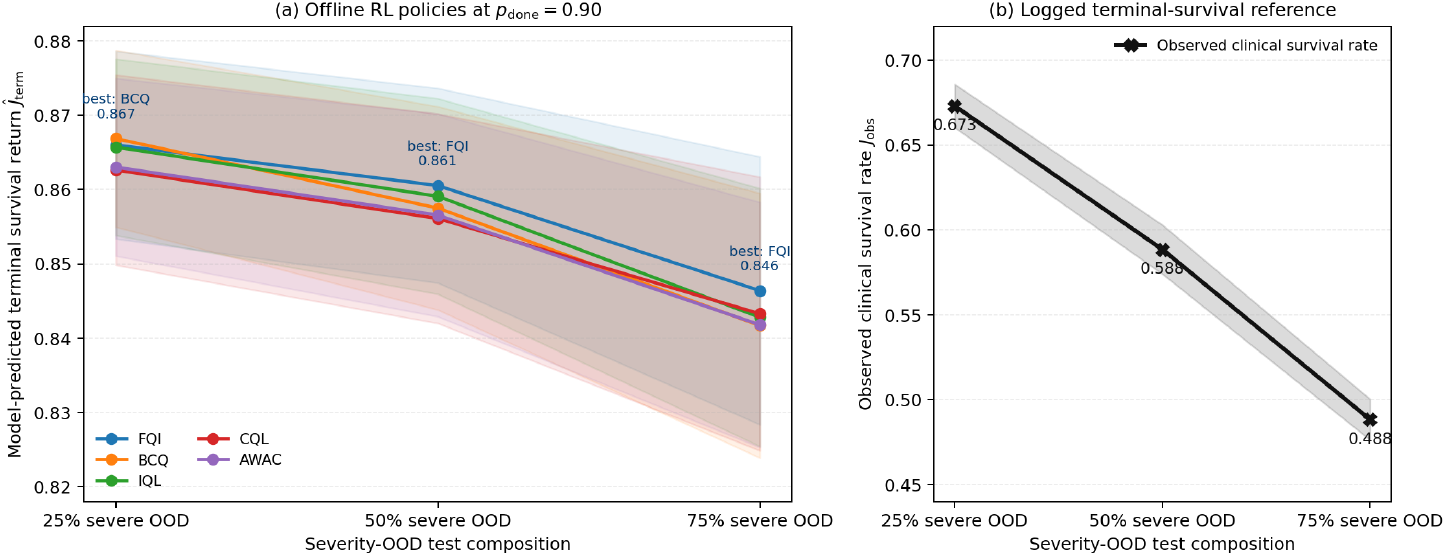
Severity-OOD main comparison at *p*_done_ = 0.9. **(a)** model-predicted terminal survival return *Ĵ*_term_ for the offline RL policies, with 95% confidence intervals across 40 seeds. The best-performing policy in each OOD setting is shown. **(b)** observed clinical survival rate *J*_obs_ for the corresponding held-out cohorts.

In contrast, the model-predicted terminal survival returns of offline RL methods remain comparatively stable across the same severity-OOD levels. Figure 1(a) illustrates this trend, where at *p*_done_ = 0.9, the average predicted returns across the three mixtures are 0.858 (FQI), 0.855 (BCQ), 0.856 (IQL), 0.854 (CQL), and 0.854 (AWAC). Accordingly, the best-performing method is BCQ at 25% severe-OOD (0.867), while FQI achieves the highest return at both 50% (0.861) and 75% (0.846) sets. Overall, the leading offline RL methods exhibit only modest degradation as cohort severity increases, suggesting a relatively stable model-based decision-support signal under this stress test. Table 3 reports the corresponding headline values at *p*_done_ = 0.9, including the observed clinical survival rate, the best model-predicted offline return, and their model-based gap.

**Table 3.** Main severity-OOD results at *p*_done_ = 0.9. The table reports the observed clinical survival rate, the best model-predicted offline return, and the model-based gap between them for each severity-OOD setting. This gap compares different quantities and should not be interpreted as a causal treatment effect.

| Method | 25% | 50% | 75% | Mean |
| --- | --- | --- | --- | --- |
| Observed | 0.673 | 0.588 | 0.488 | 0.583 |
| Best model-predicted | 0.867 | 0.861 | 0.846 | 0.858 |
| Gap: best offline - observed | +0.194 | +0.272 | +0.358 | +0.275 |
| FQI | 0.866 | **0.861** | **0.846** | 0.858 |
| BCQ | **0.867** | 0.857 | 0.842 | 0.855 |
| CQL | 0.863 | 0.856 | 0.843 | 0.854 |
| IQL | 0.866 | 0.859 | 0.843 | 0.856 |
| AWAC | 0.863 | 0.857 | 0.842 | 0.854 |

### 3.2 Sensitivity to Rollout Stopping Threshold

Figure 2 summarizes the model-predicted terminal survival returns of all methods across the severity-OOD and *p*_done_ sweep. FQI achieves the highest performance in 10 out of 12 settings, while BCQ is the best in the remaining two (25% severe-OOD at *p*_done_ *∈ {*0.7, 0.9*}*). However, the spread across offline methods is small. For instance, at 75% severe-OOD and *p*_done_ = 0.9, the evaluated methods range only from approximately 0.841 to 0.846. Therefore, the main finding is not a single dominant method, but a family of offline policies that maintain stable model-predicted terminal survival returns under severe-OOD evaluation.

**Fig. 2.**
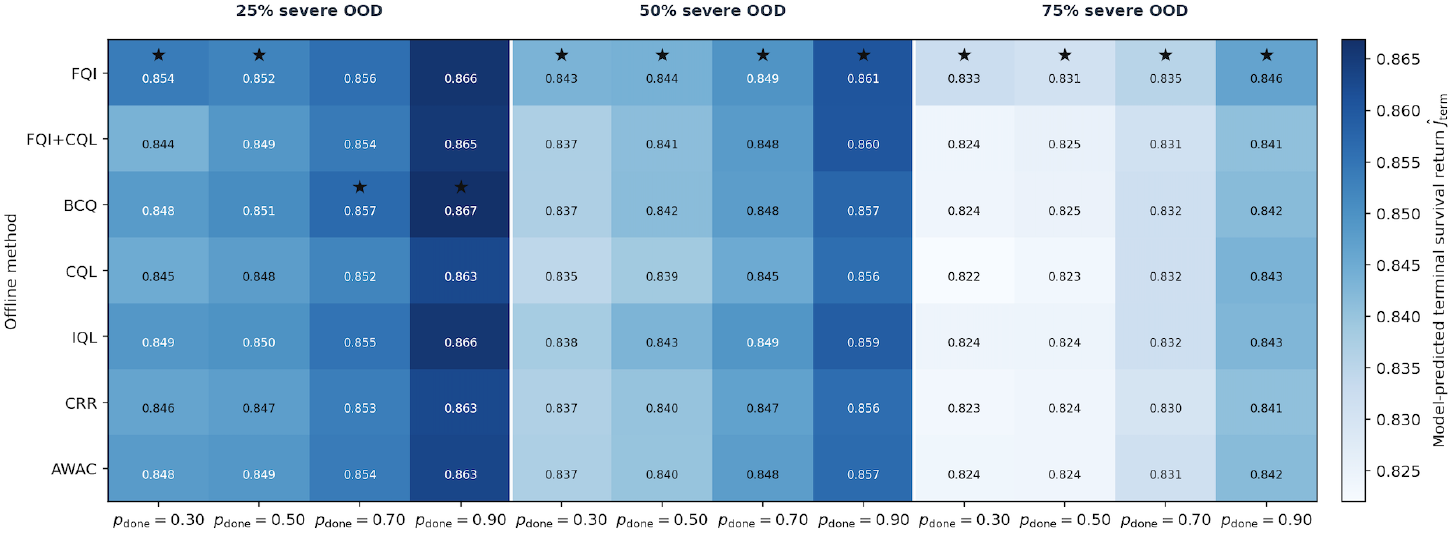
Heatmap of model-predicted terminal survival return *Ĵ*_term_ for all methods across severity-OOD ratio and *p*_done_. Stars indicate the highest-performing offline method in each setting. The figure illustrates ranking stability.

In addition, Table 4 outlines the best-offline value, the observed clinical survival rate, and their gap for every *p*_done_ across severity-OOD levels. The results demonstrate that the gap grows from *∼*0.18 at 25% severe-OOD to *∼*0.35 at 75%, an increase of about +0.16. This increase is essentially identical across all four *p*_done_ values (+0.163 to +0.164), indicating that the qualitative finding is robust to the rollout-stopping choice. This gap is a model-based contrast between distinct estimands, not a causal treatment effect.

**Table 4.** Offline-versus-observed gap across severity-OOD levels and p_done_. “Best” denotes the highest model-predicted offline return in each setting. “Observed” is the logged clinical survival rate. “Gap” is their difference. The rightmost column reports the 25%-to-75% gap increase across p_done_.

|  | 25% |  |  | 50% |  |  | 75% |  |  |  |
| --- | --- | --- | --- | --- | --- | --- | --- | --- | --- | --- |
| $p_{\text{done}}$ | Best | Observed | Gap | Best | Observed | Gap | Best | Observed | Gap | Gap incr. 25%→75% |
| 0.3 | 0.854 | 0.673 | 0.181 | 0.843 | 0.588 | 0.255 | 0.833 | 0.488 | 0.344 | +0.164 |
| 0.5 | 0.852 | 0.673 | 0.179 | 0.844 | 0.588 | 0.255 | 0.831 | 0.488 | 0.343 | +0.163 |
| 0.7 | 0.857 | 0.673 | 0.184 | 0.849 | 0.588 | 0.261 | 0.835 | 0.488 | 0.347 | +0.163 |
| 0.9 | 0.867 | 0.673 | 0.194 | 0.861 | 0.588 | 0.272 | 0.846 | 0.488 | 0.358 | +0.164 |

### 3.3 Rollout-Length Analysis

We further analyze the relationship between model-predicted terminal survival and mean rollout length for the offline methods at *p*_done_ = 0.9. Figure 3 (a) shows that mean rollout length increases with the severity-OOD ratio for all methods, while remaining similar across algorithms. Figure 3 (b) plots the corresponding scatter in the (length, *Ĵ*_term_) plane, with marker size indicating severity-OOD level. As cohort severity increases, points shift downward and to the right, reflecting lower model-predicted survival and longer rollouts. Longer rollouts are generally more susceptible to accumulated model error [8], making outcome estimates increasingly uncertain. We therefore interpret high *Ĵ*_term_ values at longer horizons as model-based decision-support signals, not as evidence of true survival improvement. Reporting rollout length alongside survival makes this explicit.

**Fig. 3.**
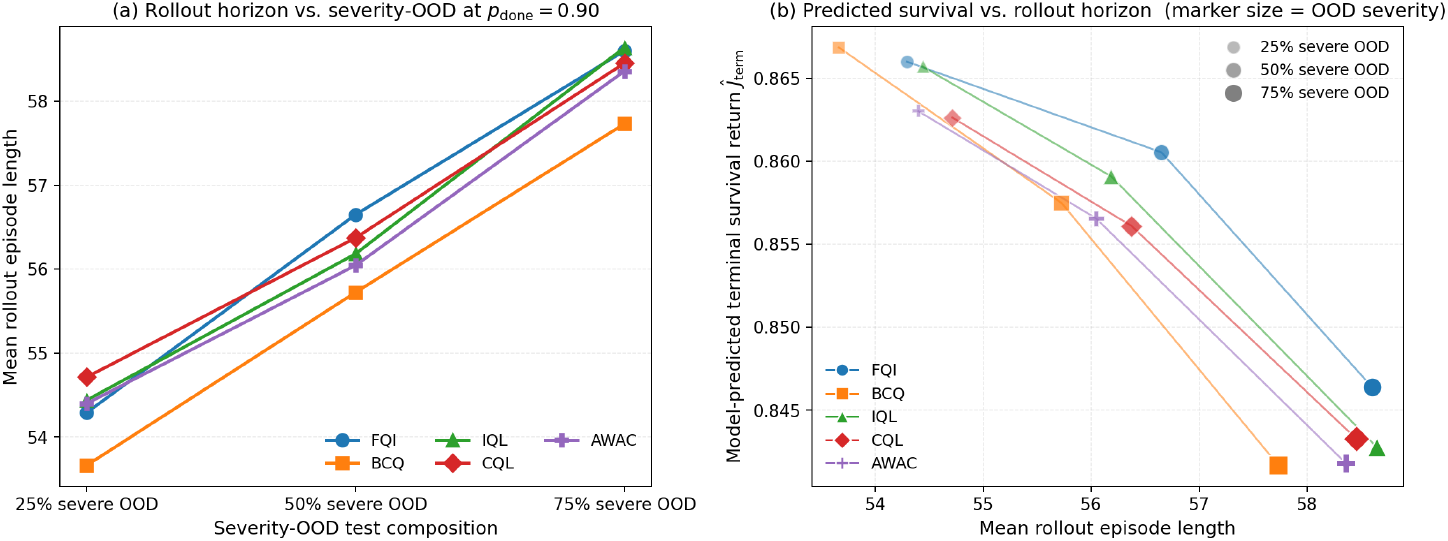
Rollout length and survival diagnostic for offline RL methods at *p*_done_ = 0.9. **(a)** Mean rollout length vs. severity-OOD ratio. **(b)** Model-predicted terminal survival return vs. mean rollout episode length, with marker size encoding severity-OOD level.

### 3.4 Physiological Stabilization Analysis

We next examine whether offline-policy model-generated rollouts exhibit more favorable physiological stabilization patterns compared to the corresponding logged clinical trajectories, as measured by the episode-level physiological stabilization score (EPSS). EPSS summarizes whether selected physiological variables improve, remain stable, or worsen over time, with higher values indicating better stabilization. Averaged across all severity-OOD levels, rollout-stopping thresholds, horizons, and random seeds, the evaluated offline methods achieve nearly identical EPSS values: 0.2078 for CQL, 0.2073 for BCQ, 0.2072 for AWAC, 0.2071 for FQI, and 0.2069 for IQL. Hence, we focus on CQL, which achieves the highest score, as a representative offline policy for the remainder of this analysis.

Figure 4 demonstrates that CQL-based model-generated rollouts at *p*_done_ = 0.9 achieve higher EPSS values than logged clinical trajectories immediately after the first rollout step and maintain this performance through horizon 20. Horizons are fixed 4-hour ICU bins, for example, *h* = 5 is approximately 20 hours after the starting point and *h* = 10 is approximately 40 hours after the initial patient state. At horizon 10, the EPSS differences are +0.23, +0.27, and +0.32 for the 25%, 50%, and 75% severe-OOD cohorts, respectively. Thus, the CQL rollouts receive higher stabilization scores than the matched observed trajectories, with the gap increasing as cohort severity grows.

**Fig. 4.**
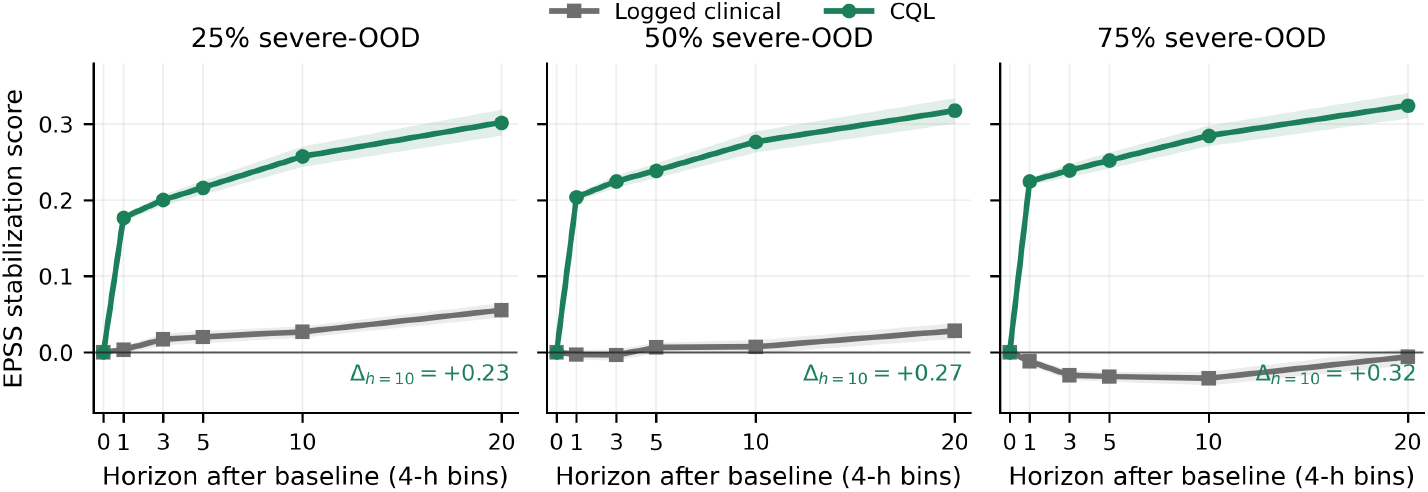
Physiological stabilization for CQL vs. logged clinical trajectories at *p*_done_ = 0.9. EPSS averages direction-corrected state-change scores over available physiological variables. Shaded intervals show 95% confidence intervals over 40 seeds.

Subsequently, we provide additional insight into the source of the EPSS differences. Figure 5 (a) presents the average CQL-minus-logged EPSS gap across the three severity-OOD mixtures, highlighting that the separation is driven primarily by the evaluation horizon rather than the rollout stopping threshold. The mean EPSS difference is approximately +0.21 at horizon 1 and reaches approximately +0.29 by horizon 20, with little dependence on *p*_done_. Figure 5 (b) further decomposes the gap into its individual physiological components for horizon 10 and *p*_done_ = 0.9. The largest positive contributions come from shock severity (+0.71), creatinine (+0.63), lactate (+0.45), and MAP (+0.32), while SpO_2_ and platelets contribute small negative differences. Overall, we attribute these findings to the broader family of offline RL techniques rather than to CQL specifically, as they all achieved nearly identical EPSS values.

**Fig. 5.**
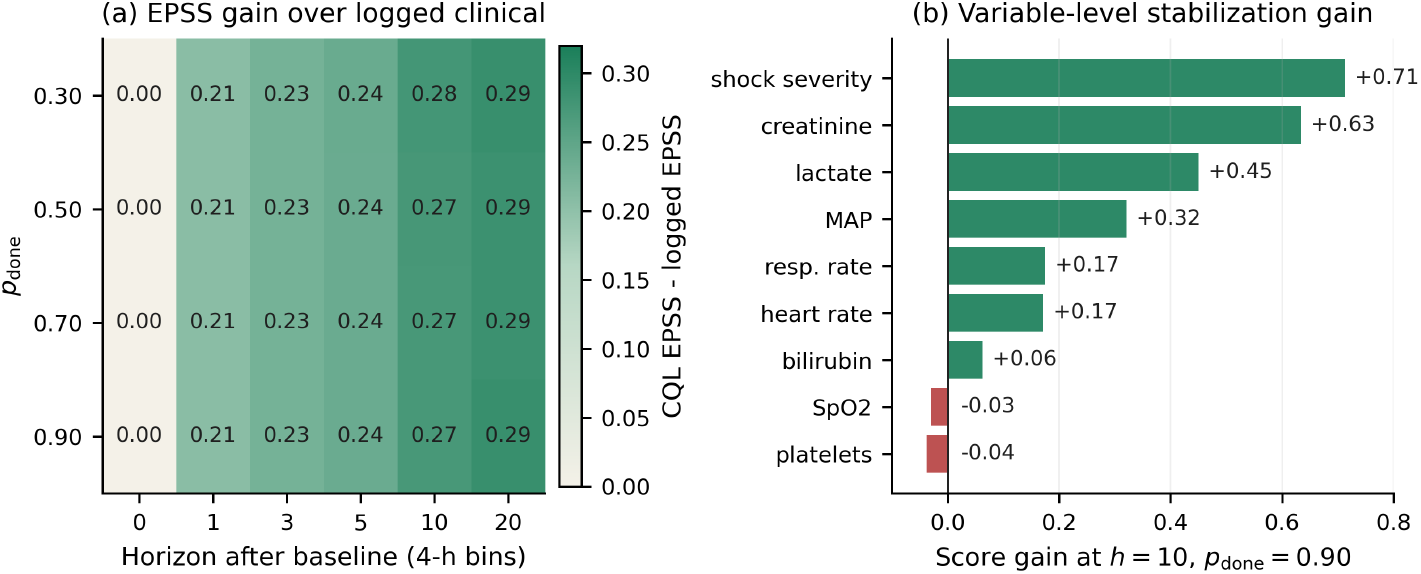
Sensitivity and component-level decomposition of the CQL physiological stabilization signal. **(a)** CQL-minus-logged EPSS averaged over severity-OOD mixtures and seeds across horizons and *p*_done_; each horizon corresponds to one 4-hour ICU bin. **(b)** Component-level EPSS difference at horizon 10 and *p*_done_ = 0.9, averaged over severity-OOD mixtures and seeds. Positive values indicate components where CQL rollouts receive higher direction-of-change scores than matched logged trajectories.

## 4 Discussion

Our experiments suggest that, under the shared learned-dynamics OPE protocol, standard offline RL policies maintain stable model-predicted terminal survival returns across all three severity-OOD mixtures and the full *p*_done_ sweep. These returns are consistently higher than the observed clinical survival rate, with the gap increasing as cohort severity grows. FQI is most consistently strongest for model-predicted terminal survival, while BCQ performs best in the mildest severe-OOD setting at longer rollouts. The physiological stabilization analysis provides a complementary signal beyond the terminal survival endpoint. In both analyses, the spread across methods is small, suggesting that the main finding is not a single winning algorithm but a family of model-based policy behavior under severity shift. These results show that under a shared learned-dynamics OPE protocol, offline RL policies exhibit stable model-based decision-support signals, but they should not be read as evidence that the learned policies would improve real patient survival or physiology.

Despite the promising results of offline RL policies under severity-induced distribution shift in ICU sepsis management, several factors should be considered. First, *Ĵ*_term_ is a model-generated OPE quantity, whereas *J*_obs_ is an observed outcome under historical clinical practice. Their difference is therefore a model-based contrast between distinct estimands, not a causal survival effect [3, 8]. Similarly, EPSS is a heuristic stabilization diagnostic computed from learned-dynamics rollouts for offline policies and from observed patient trajectories for the logged-clinical reference. As EPSS is not a validated clinical endpoint, the CQL-versus-logged gap should not be interpreted as a causal physiological effect.

Second, both *Ĵ*_term_ and policy-dependent EPSS inherit any bias or optimism in the fitted dynamics ensemble, particularly in severe-OOD states. Thus, the reported values are subject to evaluator bias which is independent of the offline policy itself. Third, the rollout-stopping threshold *p*_done_ and horizon-specific trajectory availability affect the length and composition of model-generated and logged comparisons. Longer rollouts, especially in more severe cohorts, also increase the opportunity for compounding model error (see Figure 3), a central concern in offline RL that motivates offline methods such as BCQ, CQL, and IQL in the first place [2, 7, 6]. The long-horizon EPSS comparison is also conditional on trajectory availability, with lower retained counts for logged clinical trajectories than for CQL model rollouts at *h* = 20.

Therefore, before any clinical claim or deployment consideration, future work should calibrate and validate the learned-dynamics evaluator against held-out outcomes and observed state trajectories, replicate the findings on independent clinical cohorts, and perform causal evaluation under appropriate identification assumptions. Such studies are necessary to determine whether the model-based advantages observed here translate into genuine improvements in real-world patient outcomes [3].

## 5 Conclusion

In this paper, we studied standard offline reinforcement learning methods for ICU sepsis decision support under severity-induced distribution shift. Using severity-enriched OOD test mixtures derived from the MIMIC-III benchmark, we evaluated several offline RL policies under a shared learned dynamics off-policy evaluation framework. Across all setups, the evaluated methods maintained stable model-predicted terminal survival returns, while the gap relative to the observed clinical survival rate increased as cohort severity grew. A complementary EPSS analysis revealed a consistent pattern, with offline-policy rollouts receiving higher stabilization scores than matched logged clinical trajectories. The results suggest that offline RL methods can retain stable model-based decision-support signals even in increasingly severe held-out patient cohorts. At the same time, the reported survival and physiological outcomes are model-dependent evaluation quantities and should not be interpreted as evidence of improved real-world patient outcomes. Further validation, calibration, and causal evaluation are required before clinical-use claims can be made.

## Data Availability

The data analyzed in this study were derived from the MIMIC-III Clinical Database (version 1.4), available through PhysioNet. MIMIC-III is a credentialed-access dataset, and the authors are not permitted to redistribute the underlying patient-level data. Researchers may obtain access directly from PhysioNet after completing the required credentialing, training, and Data Use Agreement.

https://physionet.org/content/mimiciii/1.4/

